# The Effect of Lifestyle-based Active Pursed-lip Diaphragmatic Breathing Training on the Prognosis of Patients with Chronic Heart Failure

**DOI:** 10.64898/2026.01.05.26343492

**Authors:** Qi Zhou, Dan Ma, Feng-jie Lv, Suxin Luo, Shenglan Yang

## Abstract

Respiratory training benefits chronic heart failure (CHF) patients but is underused due to multiple barriers. Lifestyle-based active pursed-lip diaphragmatic breathing (PLDB) offers a feasible, device-independent alternative. This study aimed to evaluate its impact on CHF patients’ exercise tolerance, cardiac function, quality of life, and clinical outcomes.

A prospective cohort study enrolled 58 hospitalized CHF patients (NYHA II-IV, Jan 2023-Feb 2024). All received standard therapy and PLDB instruction. Post-discharge, patients who self-administered PLDB were monitored via WeChat and grouped by adherence: the PLDB group (≥30 breaths/day, ≥3 days/week, n=20) and the Control group (non-compliant, n=38). Outcomes included 6-minute walk test (6MWT), grip strength, serum N-terminal pro-brain natriuretic peptide, left ventricular ejection fraction (LVEF), and quality of life indicators (Minnesota Living with Heart Failure Questionnaire [MLHFQ], Self-rating Somatic Symptom Scale-China [SSS-CN], Patient Health Questionnaire-9, Generalized Anxiety Disorder-7 [GAD-7], Pittsburgh Sleep Quality Index). Clinical endpoints were rehospitalization and major adverse cardiovascular events (MACE).

Over 6 months, the PLDB group showed significant improvements in 6MWT (333.75 ± 148.57 m to 407.9±153.29m, p=0.012), LVEF (46.67±13.86% to 51.81±14.89%, p=0.002), and quality of life (MLHFQ: 42.25±16.00 to 18.75±13.47, p<0.001; GAD-7: 3.90±3.81 to 1.50±2.14, p=0.029; SSS-CN: 35.35±5.84 to 29.05±8.61, p=0.005), with lower MACE incidence and higher MACE-free survival.

Six-month lifestyle-based active PLDB training improves exercise capacity, cardiac function, quality of life, and reduces MACE in CHF patients, serving as an effective, practical rehabilitation strategy.

## 1. Introduction

Chronic heart failure (CHF), the terminal stage of various cardiac diseases, signifies structural and functional changes in the heart, leading to a complex array of symptoms, with fatigue and dyspnea being the most prevalent. Studies reveal that 59% of CHF patients experience fatigue, while over 69% suffer from dyspnea.^1^ These symptoms not only restrict patients’ physical activity and daily functioning but also diminish their social interactions, resulting in decreased self-esteem and a significant rise in anxiety and depression. Furthermore, they serve as independent predictors of rehospitalization and mortality.^2^ Pursed-lip diaphragmatic breathing (PLDB), a respiratory training method that combines pursed-lip breathing with diaphragmatic breathing, has emerged as an effective strategy to improve cardiopulmonary function, enhance exercise tolerance, and ultimately improve patients’ quality of life and prognosis.^3,4^ Unlike traditional exercise rehabilitation, PLDB necessitates no equipment, is not constrained by location or time, and its movements are straightforward and easily learned, even by elderly patients, those with low literacy levels, and those with decompensated HF. Building upon this, Lifestyle-based Active PLDB Training offers a flexible approach, allowing patients to integrate respiratory training into their daily routines, fostering a sustainable lifestyle over the long term. This study aims to explore the impact of Lifestyle-based Active PLDB Training on cardiac function, exercise tolerance, quality of life, and prognosis in CHF patients, and identify a potentially more convenient and effective cardiac rehabilitation method for CHF patients.

## 2. Methods

### 2.1 Population

This study was conducted in the First Affiliated Hospital of Chongqing Medical University located in southwest China from January 2023 to February 2024. Inclusion Criteria: (1) Fulfilling the diagnostic criteria for chronic heart failure outlined in the “Chinese Guidelines for the Diagnosis and Treatment of Heart Failure 2018”.^5^ (2) Classified as New York Heart Association (NYHA) Functional Class II-IV. (3) Being 18 years of age or older. (4) Having voluntarily signed an informed consent form. (5) Being mentally alert and having normal communication abilities. Exclusion Criteria: (1) Having comorbidities that affect cardiopulmonary function. (2) Having severe dysfunction of the liver, kidneys, brain, or other vital organs. (3) Having a diagnosis of malignancy. (4) Having metabolic disorders, orthopedic-rheumatological or infectious diseases, or undergoing hormone or chemotherapy treatments. (5) Having any substance abuse or cognitive impairment. (6) Having a history of cardiopulmonary rehabilitation therapy or any significant non-cardiac issues that may adversely affect survival rates during the study period.

### 2.2 Group Classification and Intervention Strategies

61 patients with CHF were enrolled, and baseline data (including gender, age, body mass index [BMI], smoking history, and medication status) were recorded for each patient. All patients underwent routine anti-heart failure treatment, and their cardiopulmonary fitness, quality of life, and cardiac function were evaluated until their symptoms were alleviated. During hospitalization, patients received rehabilitation training education and instruction on correct PLDB techniques to ensure they fully understood the exercise methods. Patients were recommended to perform the exercises at least 30 times daily, with training frequency and duration determined by the patients themselves. A WeChat group was established to facilitate standardized communication and support follow-up, including monitoring patients’ training adherence and tracking their clinical status throughout the follow-up period. Patients who adhered to the PLDB protocol (performing at least 10 repetitions/set, 3 sets daily, and at least 3 days per week) were categorized as the respiratory training group, while the remaining patients were defined as the control group. Six months later, all patients underwent reassessment of exercise tolerance, muscle strength, quality of life, psychological status, and cardiac function.

### 2.3 Treatment and Respiratory Training

During hospitalization, both groups received treatment and guideline-directed medical therapy, including diuretics, recombinant human brain natriuretic peptide, levosimendan, digitalis, vasodilators, beta-blockers, angiotensin-converting enzyme inhibitors/angiotensin receptor blockers/angiotensin receptor neprilysin inhibitors, mineralocorticoid receptor antagonists, sodium-glucose cotransporter-2 inhibitors, and medications for blood pressure and blood sugar control. After discharge, patients continued to take oral heart failure medications as prescribed by their doctors. The key points of PLDB training are as follows: The patient could be in a seated or recumbent position, inhale through the nose for 3-4 seconds, then exhale slowly through pursed lips (in a kissing position) for 4-6 seconds. Repeat for 10-15 cycles per session (≥30 cycles/day) (Figure 1).

**Figure 1.**
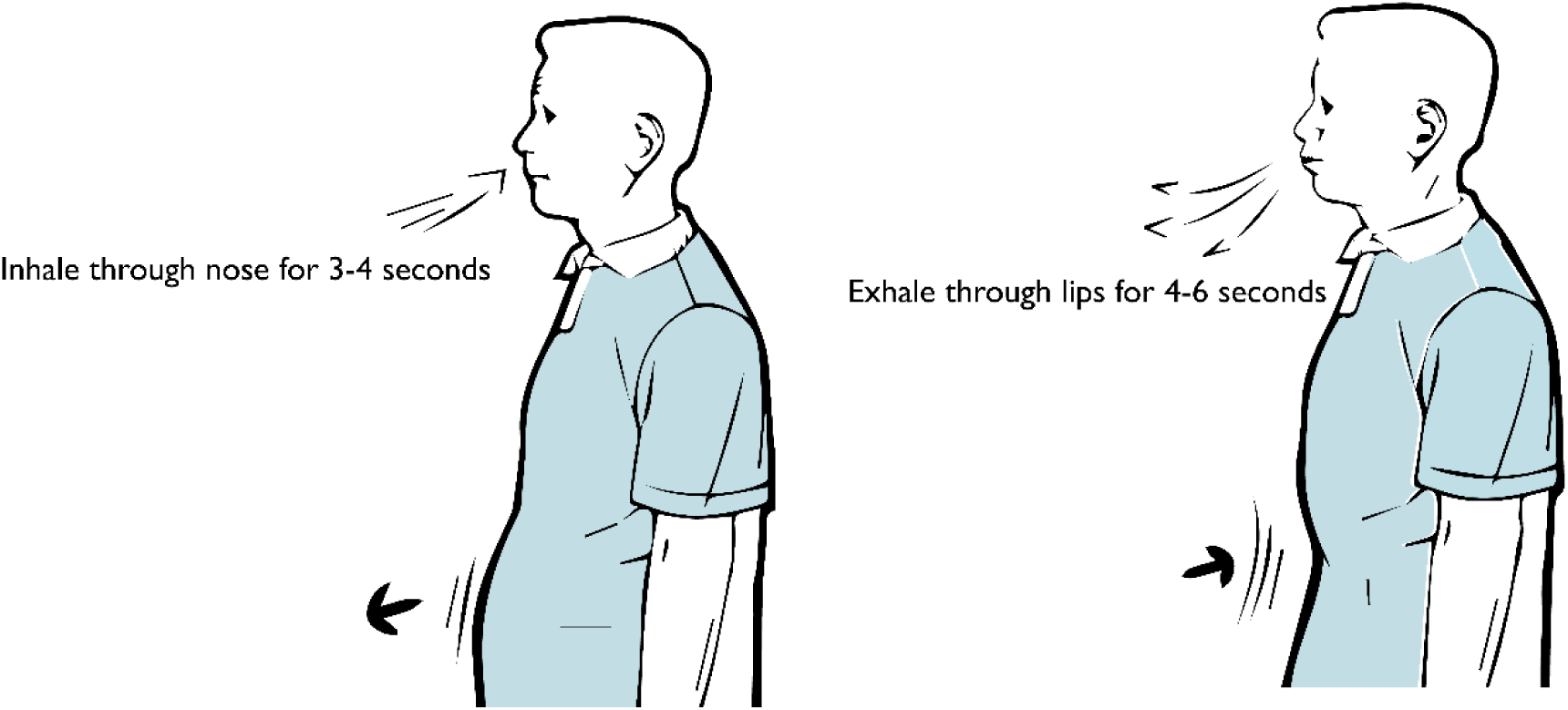
Schematic diagram of PLDB movements.

### 2.4 Trial outcomes

#### Primary outcomes

The primary outcomes were all-cause rehospitalization and major adverse cardiovascular events (MACE) at 6 months. All-cause rehospitalization is defined as a hospital stay of >24 hours, and MACE as a composite endpoint encompassing rehospitalization for acute heart failure, malignant arrhythmias, acute myocardial infarction, and cardiac death.

#### Secondary outcomes

The secondary outcomes included functional capacity, cardiac function indicators, and quality of life indicators. (1) Functional capacity indicators were assessed using the 6-Minute Walk Test (6MWT) and handgrip strength. The 6MWT was performed in a 30-meter flat, obstacle-free corridor, with patients walking back and forth at their tolerable pace to cover the maximum distance in 6 minutes. Handgrip strength was measured as follows: patients stood with feet apart and arms hanging naturally, gripped a dynamometer with their non-dominant hand, and squeezed maximally for 3-5 seconds. The test was repeated after a 15-second interval, and the maximum value was used for analysis. (2) Cardiac function indicators included serum N-terminal pro-brain natriuretic peptide (NT-proBNP) and left ventricular ejection fraction (LVEF). NT-proBNP was measured within 24 hours prior to discharge, while LVEF was determined via echocardiography using Simpson’s method. (3) Quality of life indicators were evaluated using the Minnesota Living with Heart Failure Questionnaire (MLHFQ);^6^ the Self-rating Somatic Symptom Scale-China (SSS-CN);^7^ other measures included the Patient Health Questionnaire-9 (PHQ-9),^8^ Generalized Anxiety Disorder-7 (GAD-7) scores,^9^ and Pittsburgh Sleep Quality Index (PSQI).^10^ Higher scores indicate poorer quality of life.

#### Follow-up

Follow-up for each patient commenced upon discharge and lasted 6 months, with real-time adherence tracking supported by WeChat monitoring and the hospital’s electronic medical record system. The follow-up concluded at the earliest of two events: completion of the predetermined 6-month duration or occurrence of MACE. Throughout follow-up, all-cause rehospitalization and MACE were recorded in detail. Patients were instructed to recomplete assessments of all outcome indicators upon completion of the follow-up period.

### 2.5 Statistical Analyses

All continuous variables were expressed as mean ±standard deviation, and non-normally distributed variables as median (interquartile range). Baseline comparisons between groups were performed using Student’s t-test for normally distributed variables (age, BMI, 6MWT, hand grip strength, MLHFQ, GAD-7, PHQ-9, PSQI, SSS-CN, and LVEF). For non-normally distributed variables (NT-proBNP), the Mann-Whitney U test was utilized. Nominal and categorical variables (gender, smoking history, medication history, and prognostic indicators) were assessed using Fisher’s exact test and the Chi-squared test. The normal distribution of data was assessed using the Kolmogorov-Smirnov test. Wilcoxon’s signed-rank test and paired t-test were used to assess training-induced changes (before and after) within a particular group. For survival analysis, the Kaplan-Meier method was used to construct MACE-free survival curves, with differences between groups compared using the log-rank test. Follow-up time was calculated from discharge (initiation of PLDB training) to the first occurrence of MACE (considered an endpoint event) or the end of the 6-month period. Patients without MACE by the 6-month mark were censored at the final follow-up assessment. p-values<0.05 were considered statistically significant for all tests. Data were analyzed using SPSS 22.0, and graphs were plotted using GraphPad Prism 9.5.

### 2.6 Ethics considerations

This study was approved by the Medical Ethics Committee of the First Affiliated Hospital of Chongqing Medical University and was performed in accordance with the Declaration of Helsinki. Participants were notified concerning the training and its possible risks. Informed consent was obtained from all the participating patients.

## 3. Results

### 3.1 Baseline Characteristics of the Study Population

61 patients were initially enrolled in the study. Among them, 1 patient was excluded due to severe infection during hospitalization, 1 due to a subsequent diagnosis of systemic lupus erythematosus, and 1 due to self-discontinuation of medication after discharge. Therefore, a total of 58 patients completed the 6-month follow-up. Based on their adherence to respiratory training after discharge, patients were divided into the respiratory training group (n=20) and the control group (n=38) (Figure S1). No significant differences were observed between the two groups in terms of baseline demographic details, including age, gender, BMI, smoking history, and medications (Table 1).

**Table 1.**
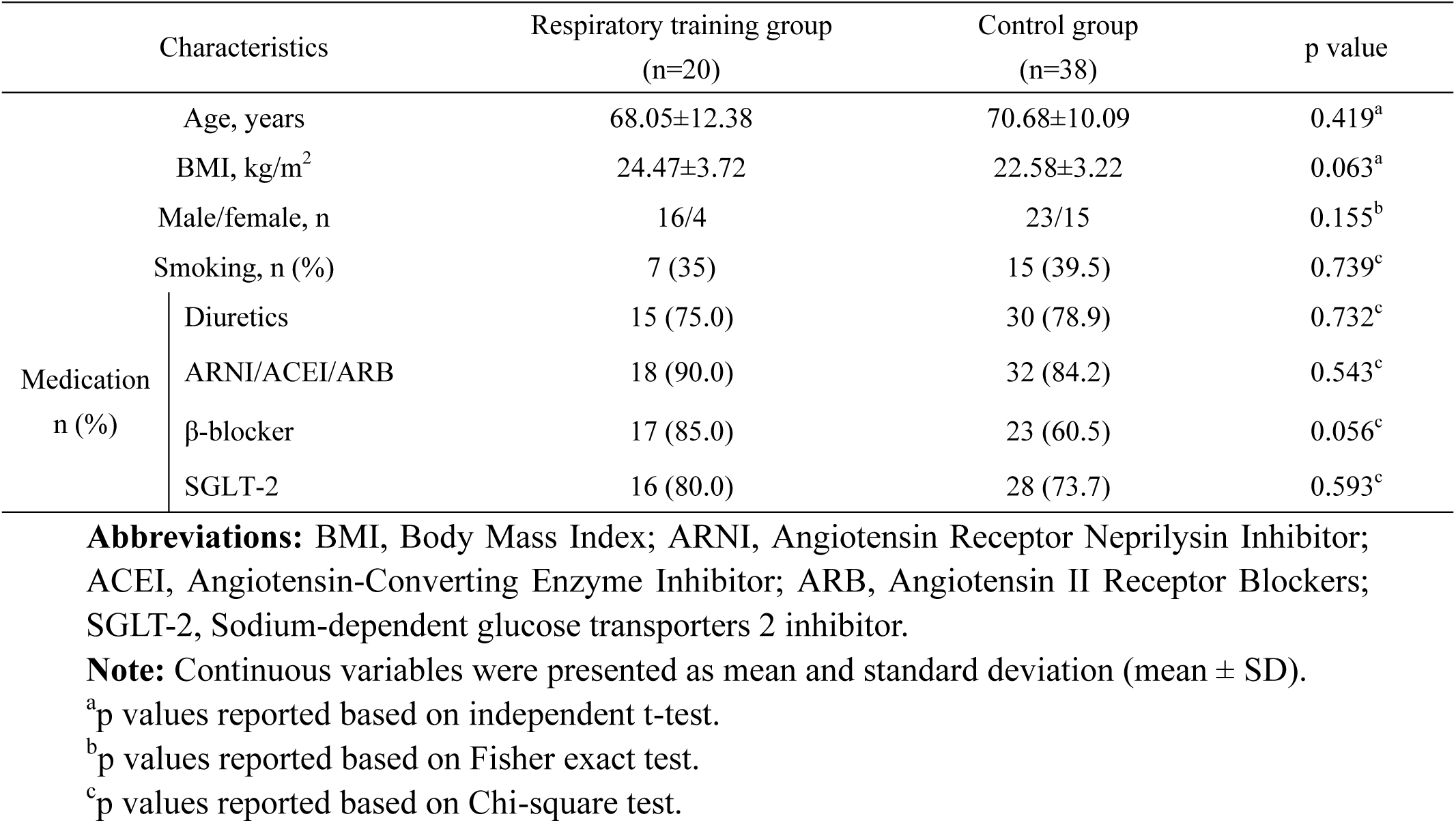
Baseline demographic characteristics of respiratory training and control groups.

There was no statistically significant difference in 6MWT distance and hand grip strength between the two groups before intervention. After 6 months, the 6MWT distance in the respiratory training group showed a significant increase (333.75±148.57 m to 407.9±153.29 m, p=0.012). In the control group, however, the 6MWT distance showed a declining trend after 6 months (381.86±131.71 m to 367.98±141.39 m), but the difference was not statistically significant (p=0.379, Figure S2). In intra-group comparisons, no statistically significant change in hand grip strength was observed over the 6-month follow-up period in either the respiratory training group or the control group (Table 2).

**Table 2.**
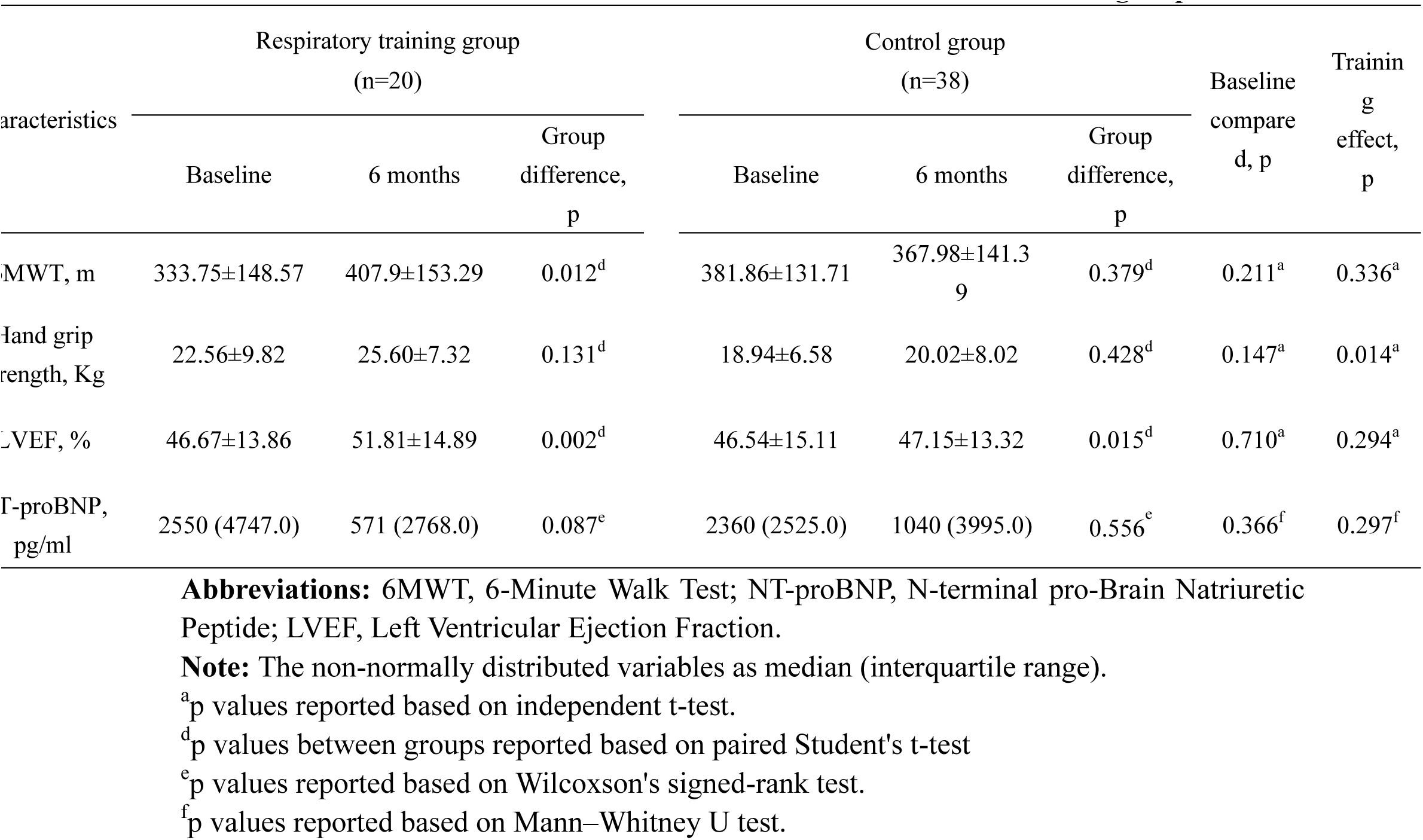
Exercise tolerance and cardiac function between the two groups.

There was no statistically significant difference in LVEF between the two groups before intervention (46.67 ± 13.86% vs 46.54 ± 15.11%, p=0.211). After 6 months, LVEF in both the respiratory training group and the control group increased significantly compared with baseline (respiratory training group: 46.67±13.86% to 51.81±14.89%, p=0.002; control group: 46.54±15.11% to 47.15±13.32%, p=0.015) (Table 4, Figure S3). LVEF in the respiratory training group was higher than that in the control group, but no significant difference was attained (51.81±14.89% vs 47.15± 13.32%, p=0.294). There was no statistically significant difference in NT-proBNP between the two groups at baseline or post-intervention, nor within either group, although the respiratory training group showed a decreasing trend (Table 4).

**Table 3.**
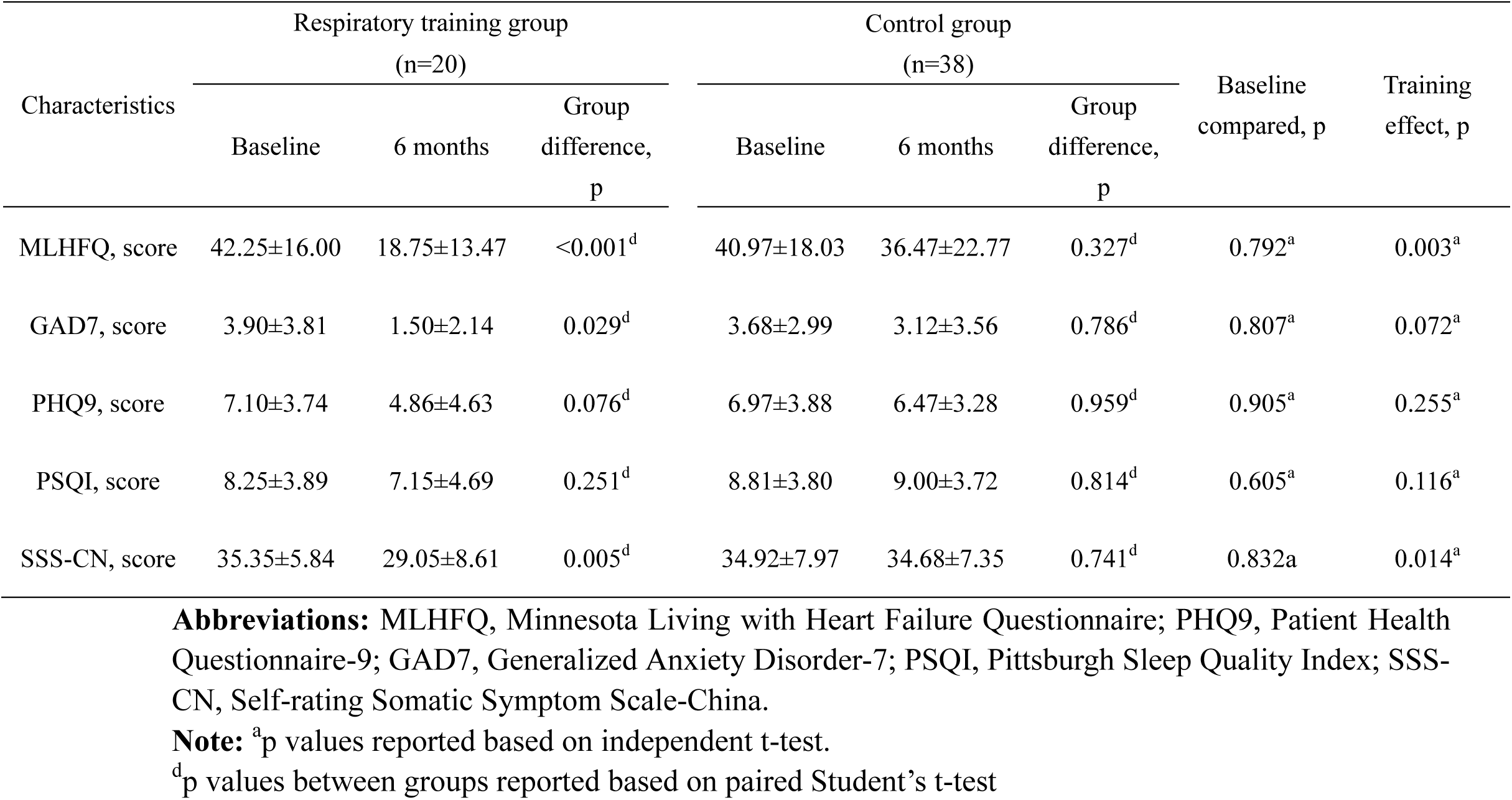
Quality of life indicators between the two groups.

**Table 4.**
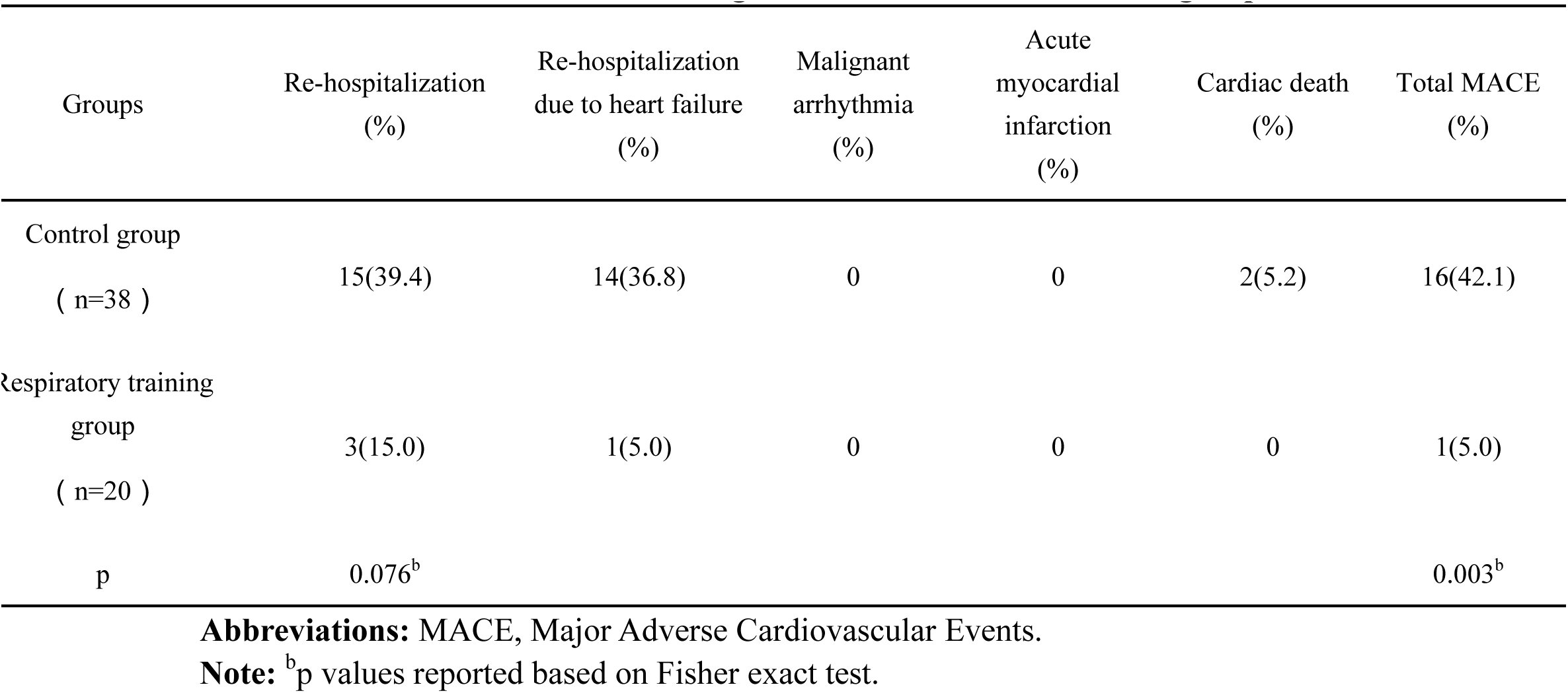
The 6-Month Prognostic Events between the two groups.

### 3.3 Improvements in Quality of Life and Mental Status

The between-group analysis showed no statistically significant differences in MLHFQ, GAD-7, PHQ-9, PSQI, and SSS-CN scores before the intervention. Six months later, the MLHFQ and GAD-7 scores in the respiratory training group were significantly decreased compared with the pre-intervention levels (MLHFQ: 42.25±16.00 to 18.75±13.47, p < 0.001; GAD-7: 3.90±3.81 to 1.50 ±2.14, p = 0.029; SSS-CN: 35.35±5.84 to 29.05±8.61, p=0.005). Additionally, the MLHFQ and SSS-CN scores in the respiratory training group were lower than those in the control group after intervention (MLHFQ: 18.75±13.47 vs 36.47±22.77, p=0.003; SSS-CN: 29.05±8.61 vs 34.68±7.35, p=0.014) (Table 3), suggesting a better quality of life and psychological health after respiratory training.

### 3.4 Adverse Clinical Events and Kaplan-Meier Survival Analysis for MACE-Free Survival

During the follow-up period, there were 3 rehospitalization events in the respiratory training group, including 1 due to recurrence of decompensated heart failure. In contrast, the control group experienced 15 rehospitalizations, with 14 of these being attributed to worsening heart failure. No cases of cardiogenic death were observed in the respiratory training group, whereas 2 cardiogenic deaths occurred in the control group. Hence, the incidence of composite MACE events was significantly lower in the respiratory training group compared with the control group (5% vs 42.1%, p=0.003, Table 5).

Kaplan-Meier survival analysis revealed a significantly higher MACE-free survival rate in the PLDB group compared with the control group (log-rank p=0.004, Figure 2).

**Figure 2.**
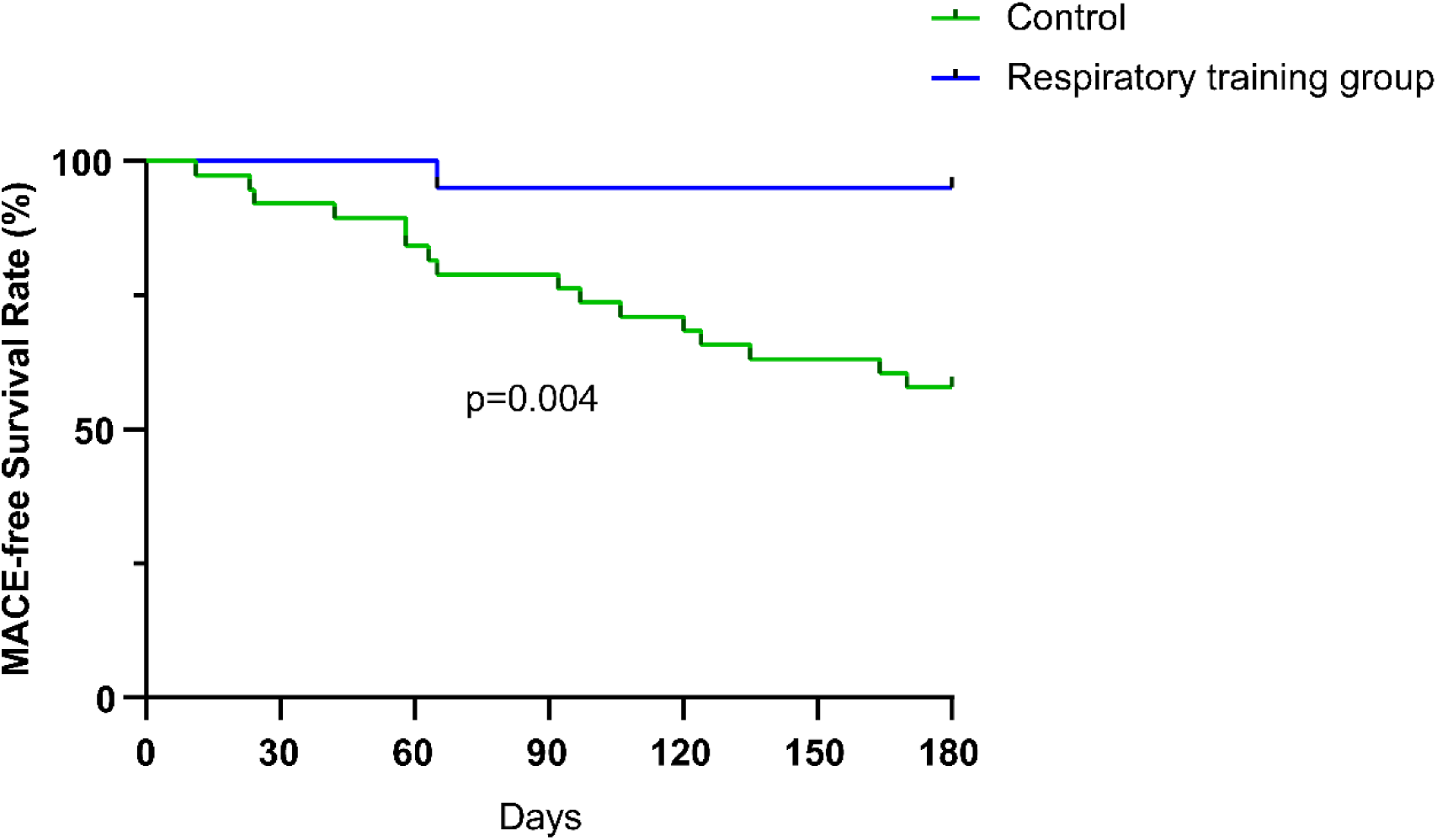
Kaplan-Meier survival analysis of MACE-free survival rate between the two groups. Abbreviations: MACE, Major Adverse Cardiovascular Events.

## 4. Conclusion

The present findings showed that CHF patients who underwent 6-month Lifestyle-based Active PLDB Training experienced significant improvements in physical fitness and showed an improving trend in cardiac function. Furthermore, their quality of life and sleep quality improved, with relief in anxiety symptoms. Notably, during the 6-month period, there was a meaningful decrease in the occurrence of MACE, although the decreasing trend in rehospitalization was not statistically significantly different from that in the control group. Ultimately, these findings suggest that Lifestyle-based Active PLDB Training could be an effective, practical rehabilitation strategy for CHF patients, leading to improved clinical prognosis.

## 5. Discussion

CHF patients often suffer from dyspnea during physical activity, which is a classical feature of reduced cardiac function. The core mechanisms involve muscle atrophy due to insufficient perfusion of exercising muscles, respiratory muscle weakness, pulmonary congestion, a and sympathetic nerve dysfunction,^11–13^ which form a vicious cycle of “activity limitation-physical capacity decline-symptom exacerbation”.

Targeted inspiratory muscle training (IMT) or respiratory rehabilitation enhances inspiratory duration, reduces cardiac afterload, increases cardiac output, and improves respiratory muscle function and lung capacity.^14^ While recent guidelines have integrated such training into CHF rehabilitation protocols,^15,16^ they lack specific implementation details, making optimal respiratory training a research focus. Sadek et al. conducted a meta-analysis that identified the optimal IMT protocol: 6 sessions/week at 60% maximal inspiratory pressure (PImax) for 30 minutes over 12 weeks, which best improves inspiratory strength, 6-minute walking distance, and dyspnea.^17^ However, this approach requires specialized equipment and professional supervision, limiting its use in remote or resource-limited areas. Frail patients may also struggle to reach target intensities, increasing the risk of adverse events. Kawauchi et al. noted that low-to-moderate intensity IMT similarly improves muscle strength and exercise tolerance but shows inconsistent effects on heart failure symptoms and NYHA classification.^18^ Our study thus explores respiratory rehabilitation strategies suitable for long-term home-based practice to enhance the quality of life and prognosis of CHF patients.

The PLDB training mode shares similarities with the concept of “qi sinking to the dantian” in Tai Chi, a traditional Chinese exercise, emphasizing the regulation of physical and mental states through slow and continuous breathing rhythms. Our study demonstrated that PLDB can effectively enhance 6MWT performance, improve cardiac function and quality of life, and alleviate anxiety in CHF patients, while also reducing the incidence of MACE events within 6 months. Possible explanations for these findings include: (1) Pursed-lip exhalation increases intratracheal pressure and reduces small airway collapse; diaphragmatic breathing strengthens diaphragmatic movement and lowers respiratory resistance.^19,20^ These two effects synergistically reduce cardiac preload. Furthermore, diaphragmatic breathing promotes coordinated abdominal muscle contraction, effectively improving the coordination and activity of respiratory muscles, such as the abdominal muscles, internal intercostal muscles, diaphragm, and lower chest muscles, which tend to be inefficient when HF congestion occurs, thereby significantly enhancing pulmonary ventilation dynamics. The improvement in pulmonary ventilation further optimizes the aerobic metabolism of skeletal muscles, promotes muscle fiber growth and muscle protein synthesis, improves muscle adaptability, and enhances exercise tolerance.^21^ Together, these factors ultimately help break the vicious cycle of “activity limitation - physical capacity decline - symptom exacerbation” (Figure S4).

(2) Slow-deep breathing can enhance parasympathetic tone, inhibit the sympathetic nerve, improve heart rate variability, and reduce norepinephrine levels, thereby decreasing myocardial oxygen consumption and the risk of arrhythmias.^22–24^ Following sympathetic inhibition, the activity of aldosterone and the renin-angiotensin system is further suppressed, reducing the release of cardiotoxic mediators and alleviating myocardial injury.^2,25^ This may be the key to the reduction of MACE events. (3) Improvements in cardiopulmonary function alleviate fatigue and dyspnea, thereby enhancing patients’ daily autonomy and self-management capacity. Such physiological improvements were associated with reduced anxiety and insomnia symptoms, accompanied by improved quality of life. These psychophysiological benefits are reproducible across clinical populations, as validated in both COPD and cancer patient cohorts.^26,27^

Enhancing the adherence and applicability of cardiac rehabilitation training has long been a focus of research. Islam et al. found that “exercise snacks”——defined as multiple brief bouts of vigorous exercise performed throughout the day——can promote cardiovascular health.^28^ Stamatakis et al. demonstrated that incidental physical activity (IPA), such as active transport, household activities, or work-related interventions, can reduce the risk of cardiovascular disease (CVD). For individuals unable to sustain long-term structured exercise, IPA serves as an effective means of preventing CVD.^29^ This indicates a gradual shift in rehabilitation protocols from structured, long-duration models toward fragmented, lifestyle-integrated approaches, emphasizing transformations in lifestyle and rehabilitation concepts. In our study, we provided participants with reference minimum weekly training goals, enabling them to engage in daily training flexibly according to their individual lifestyles; we term this approach “lifestyle-based active training”. Its advantages include higher safety (no risk of exercise-induced ischemia), broader applicability (tolerable even for CHF patients with NYHA IV), and lower cost (no need for specialized equipment). Unlike traditional exercise methods, which are constrained by venue and intensity limitations, lifestyle-integrated breathing training can be incorporated into daily routines (e.g., after waking up or meals), making it easier to sustain over the long term.

Several limitations merit consideration in interpreting these findings: (1) As a single-center study with a relatively modest sample size (n = 58), the generalizability of results may be constrained by local practice patterns and reduced statistical power, which may limit subgroup analyses. (2) As PLDB training requires sustained patient engagement (≥30 cycles/day), the open-label design may have introduced recruitment bias by disproportionately attracting patients with greater self-efficacy or positive attitudes toward non-pharmacologic therapies, as observed in comparable rehabilitation trials.

(3) While the 6-month follow-up demonstrated promising effects on physiological and quality-of-life endpoints, extended observation would be needed to evaluate lasting impacts on major clinical outcomes. (4) Additionally, the optimal respiratory training parameters (frequency/duration/breathing depth) for PLDB remain undefined, and unmeasured confounders, such as baseline pulmonary function, may contribute to residual bias. These limitations underscore the need for confirmatory multicenter trials with extended follow-up periods.

## Acknowledgements

This work was financially supported by the Scientific Research Project of Chongqing Municipal Sports Bureau (Project No. B202465), which is gratefully acknowledged. The authors would like to express their sincere thanks to Professor Shenglan Yang and Suxin Luo for her valuable guidance on the study design and manuscript revision. Additionally, the authors appreciate the participation of all subjects involved in this study.

## 9. Authors’ Contributions

Qi Zhou: Participated in the design of the study, guided respiratory training, conducted patient follow-up, drafted the initial manuscript, performed data statistical analysis, coordinated team meetings, and summarized intermediate research progress; Dan Ma, Feng-jie Lv: Collaborated on data entry, assisted in patient follow-up, and participated in data quality control (cross-checking data); Shenglan Yang, Suxin Luo: Led the formulation of the research protocol (methodology design, patient inclusion/exclusion criteria), revised the manuscript, and jointly completed the ethical approval application. All authors have read and approved the final manuscript and agree with the author order.

## 10. Data availability

The data underlying this article cannot be shared publicly due to the privacy of individuals that participated in the study. The data will be shared on reasonable request to the corresponding author.

## Abbreviations

BMI: Body Mass Index
CHF: Chronic Heart Failure
CVD: Cardiovascular Disease
GAD-7: Generalized Anxiety Disorder-7
IMT: Inspiratory Muscle Training
IPA: Incidental Physical Activity
LVEF: Left Ventricular Ejection Fraction
MACE: Major Adverse Cardiovascular Events
MLHFQ: Minnesota Living with Heart Failure Questionnaire
6MWT: 6-Minute Walk Test
NT-proBNP: N-terminal pro-Brain Natriuretic Peptide
NYHA: New York Heart Association
PHQ-9: Patient Health Questionnaire-9
PLDB: Pursed-Lip Diaphragmatic Breathing
PSQI: Pittsburgh Sleep Quality Index
SSS-CN: Self-rating Somatic Symptom Scale-China

